# Speed and accuracy of whole-genome nanopore sequencing for differential *Neisseria gonorrhoeae* strain detection in samples collected prospectively from a sexual health clinic

**DOI:** 10.1101/2022.04.24.22272897

**Authors:** LT Phillips, AA Witney, M Furegato, KG Laing, L Zhou, ST Sadiq

## Abstract

**Background:** Antimicrobial resistance (AMR) in *Neisseria gonorrhoeae* is a continuing global health challenge. Limitations to current national surveillance systems for reporting AMR trends, alongside reduction in culture-based diagnostics and susceptibility testing, has led to an increasing need for rapid diagnostics and identification of circulating *N. gonorrhoeae* strains. We investigated nanopore based sequencing time and depth needed to accurately identify closely related *N. gonorrhoeae* isolates, compared to Illumina MiSeq sequencing.

**Methods:** *N. gonorrhoeae* strains prospectively collected from a London Sexual Health clinic were sequenced on both Illumina MiSeq and Oxford Nanopore Technologies (ONT) MinION platforms. Accuracy was determined by comparing variant calls at 68 nucleotide positions representing 37 pre-characterised resistance associated markers in *N. gonorrhoeae*. Accuracy at varying MinION sequencing depths were determined through retrospective analysis of time-stamped reads.

**Results:** Of the 22 MinION-MiSeq sequence pairs that reached sufficient depth of coverage for comparison, overall agreement of variant call positions passing quality control criteria was 185/185 (95% CI: 98.0-100.0), 502/503 (95% CI: 98.9-99.9) and 564/565 (95% CI: 99.0-100.0) at 10x, 30x and 40x MinION depth, respectively. Isolates found to be genetically closely related by MiSeq, that is within one yearly evolutionary distance of ≤5 single nucleotide polymorphisms, were accurately identified as such via MinION.

**Conclusion:** Nanopore based sequencing shows utility for use as a rapid surveillance tool to correctly detect closely related *N. gonorrhoeae* strains, with just 10x sequencing depth, taking a median sequencing time of 29 minutes. This highlights its potential utility for tracking local gonorrhoea transmission and AMR markers.

## Introduction

The spread of *Neisseria gonorrhoeae*, resistant to multiple antimicrobial classes, has created a major global health challenge [1-4]. Effective antimicrobial resistance (AMR) phenotypic surveillance programs provide early indication of AMR spread and inform treatment guidelines [4, 5]. Although for most antibiotic classes *N. gonorrhoeae* AMR prevalence exceeds the conventional 5% threshold for abandoning empirical use, variable numbers remain susceptible to these classes, depending on geographical region. In the 2019-20 Gonococcal Resistance to Antimicrobials Surveillance Programme (GRASP) in England 55.7% *N. gonorrhoeae* strains were susceptible to ciprofloxacin [5] compared to >90% in four countries in Southeast Asia [4].

Directed *N. gonorrhoeae* treatment, enabled by culture and phenotypic susceptibility testing, has long turn-around times and has been largely replaced with nucleic acid amplification tests for diagnosis, potentially impacting the ability of surveillance programs to disrupt AMR spread [6]. Molecular *N. gonorrhoeae* AMR genotypic markers correlate variably with phenotypic resistance due to the complexity of genetic based resistance [7, 8]. However, some molecular markers are potentially useful in laboratory-based [9] or point-of-care tests [10-13]. For ciprofloxacin resistance, molecular AMR markers have remained relatively unchanged [14-16], associated mainly with the quinolone resistance determining region of the *gyrA* gene. For macrolides, penicillins and tetracyclines, diverse mechanisms and continual evolution suggests molecular markers may only be useful in combination with predictive modelling tools [16].

Recent studies suggest whole genome sequencing (WGS) from *N. gonorrhoeae* cultures has utility for molecular surveillance [17-20] and AMR prediction [], highlighting collectively that the number of single nucleotide polymorphism (SNP) expected to contain all direct and indirect transmission pairs in samples collected a year apart may diverge by up to 14 SNPs, and collected within the same day, between 0-9 SNPs [17]. For this method to progress towards real-time surveillance, WGS will be required directly from extracted clinical samples, which has been achieved in previous *N. gonorrhoeae* WGS studies for tracking AMR in high-risk sexual networks [20, 22]. Furthermore, ‘real time’ surveillance, that could support partner notification or contact tracing might be deployable near clinical services using sequencing platforms that provide the flexibility to sequence to a requisite depth [23]. Oxford Nanopore Technologies’ (ONT) MinION device has potential to be deployed ‘near-clinic’ and be re-useable, thus effective in achieving these aims. Recent iterations of the technology have improved sequencing accuracy [24], and previous work demonstrated urinary and respiratory tract infection diagnosis and AMR detection from clinical samples within a four-hour time frame [25, 26]. Recently, following 20 hours of sequencing clinical *Staphylococcus aureas* isolates on one flow-cell, accurate sequence-type was obtained within 20 minutes, suggesting near-clinic applications are achievable if accuracy can be maintained [27].

We assessed the earliest time to accurately detect AMR variants and identify phylogenetically related *N. gonorrhoeae* isolates using MinION, as a first step towards an integrated rapid diagnostic and AMR surveillance tool.

## Methods

### N. gonorrhoeae isolates

*N. gonorrhoeae* isolates (n=58) consecutively collected between July and September 2013 from St George’s University Hospitals NHS Foundation Trust, as part of 2013 GRASP (UK Health Security Agency, formerly known as Public Health England), were retrieved from frozen growth in glycerol stocks at the Sexually Transmitted Bacteria Reference Unit, and sent back to St George’s, University of London. All cultures had undergone phenotyping for antimicrobial susceptibility as part of GRASP, using European Committee on Antimicrobial Susceptibility Testing (EUCAST) breakpoints for minimum inhibitory concentration (MIC) (Table 1) [28].

**Table 1.** *Neisseria gonorrhoeae* isolate MIC data. Minimum inhibitory concentrations for 45 *Neisseria gonorrhoeae* isolates. Quinolone MIC breakpoints defined as; susceptible ≤0.03 mg/l; intermediate <0.03 - ≤0.06 mg/l; and resistant >0.06mg/l. Macrolide MIC breakpoints defined as; susceptible ≤0.25 mg/l; intermediate <0.25 - ≤0.5 mg/l; and resistant >0.5 mg/l. Penicillin MIC breakpoints defined as; susceptible ≤0.06 mg/l; intermediate <0.07 - ≤1 mg/l; and resistant <1 mg/l. Ceftriaxone and cefixime MIC breakpoints defined as; susceptible ≤0.125 mg/l; and resistant >0.125. Red: resistant; yellow: intermediate; green: susceptible, according to EUCAST MIC breakpoints.

| Isolate ID | Quinolone MIC (mg/L) | Macrolide MIC (mg/L) | Penicillin MIC (mg/L) | Ceftriaxone MIC (mg/L) | Cefixime MIC (mg/L) |
| --- | --- | --- | --- | --- | --- |
| NG002 | 0.03 | 0.13 | 0.25 | 0.004 | 0.008 |
| NG003 | 16 | 0.5 | 1 | 0.03 | 0.125 |
| NG004 | 0.03 | 0.125 | 0.5 | 0.015 | 0.008 |
| NG005 | >16 | 0.25 | 2 | 0.03 | 0.125 |
| NG006 | 0.03 | 0.06 | 0.125 | 0.004 | 0.008 |
| NG007 | 16 | 1 | 1 | 0.06 | 0.06 |
| NG008 | >16 | 0.25 | 1 | 0.015 | 0.015 |
| NG010 | 0.03 | 0.25 | 1 | 0.008 | 0.015 |
| NG011 | 8 | 0.125 | 0.25 | 0.015 | 0.06 |
| NG012 | 0.03 | 0.125 | 0.25 | 0.004 | 0.008 |
| NG013 | 0.03 | 0.125 | 0.125 | 0.004 | 0.008 |
| NG014 | 16 | 0.25 | 0.5 | 0.03 | 0.125 |
| NG015 | 0.03 | 0.125 | 0.125 | 0.004 | 0.008 |
| NG016 | 16 | 0.25 | 1 | 0.03 | 0.125 |
| NG017 | 0.03 | 0.125 | 0.25 | 0.004 | 0.008 |
| NG018 | 0.03 | 0.25 | 0.25 | 0.004 | 0.008 |
| NG019 | >16 | 0.25 | 1 | 0.008 | 0.008 |
| NG020 | 0.03 | 0.125 | 0.25 | 0.004 | 0.008 |
| NG022 | 16 | 0.5 | 0.5 | 0.03 | 0.06 |
| NG023 | 16 | 0.25 | 1 | 0.03 | 0.125 |
| NG024 | 0.03 | 0.06 | 0.25 | 0.008 | 0.06 |
| NG026 | 0.03 | 0.25 | 0.25 | 0.008 | 0.008 |
| NG027 | 0.03 | 0.125 | 0.125 | 0.008 | 0.008 |
| NG028 | 0.03 | 0.03 | 0.125 | 0.004 | 0.008 |
| NG029 | 0.03 | 0.125 | 0.25 | 0.004 | 0.008 |
| NG031 | 0.03 | 0.25 | 0.25 | ≤0.002 | 0.004 |
| NG032 | 0.03 | 0.03 | 0.125 | ≤0.002 | ≤0.002 |
| NG033 | 0.03 | 0.125 | 0.25 | ≤0.002 | 0.008 |
| NG035 | 0.03 | 0.25 | 0.25 | 0.004 | 0.008 |
| NG037 | 8 | 0.125 | 0.5 | 0.015 | 0.06 |
| NG038 | 8 | 0.125 | 0.5 | 0.015 | 0.06 |
| NG039 | 0.03 | 0.06 | 0.25 | ≤0.002 | 0.008 |
| NG040 | 0.03 | 0.06 | 0.125 | 0.004 | 0.008 |
| NG043 | 0.03 | 0.125 | 0.25 | 0.004 | 0.008 |
| NG044 | 0.03 | 0.03 | ≤0.060 | 0.004 | 0.015 |
| NG045 | 0.03 | 0.125 | 0.25 | 0.004 | 0.008 |
| NG047 | 0.03 | 0.125 | 0.125 | ≤0.002 | 0.008 |
| NG048 | 0.03 | 0.03 | ≤0.060 | ≤0.002 | ≤0.002 |

### MiSeq library preparation and sequencing

All 58 isolates were inoculated onto CBA (Oxoid, UK) plates and incubated for 24-48 hours in 5% carbon dioxide at 35°C. A 10µL loop of bacterial growth was used for genomic DNA extraction using QIAmp DNA Mini Kit (QIAGEN, UK) according to manufacturer’s instructions. Sequence libraries were prepared using Nextera XT library preparation kit (Illumina, UK) according to manufacturer’s instructions, and sequenced using paired end 2×300bp version three chemistry [29] on the MiSeq (Illumina, UK).

### MinION library preparation and sequencing

For MinION sequencing, 47/58 isolates were successfully re-grown as detailed above, DNA extraction carried out using FastDNA SPIN kit for Soil (MP Biomedicals, USA) according to manufacturer’s instructions and quantified using the Nanodrop 1000 (ThermoFisher, USA).

See supplementary material for library preparation and sequencing methods.

### Sequence alignment and variant calling

Sequence reads from both sequencing platforms were mapped to the FA1090 reference genome (RefSeq accession: NC_002946) using bwa mem (v0.7.3a-r367), alignments sorted, duplicates removed with samtools and bcftools (v1.3.1) [30], and variant calling performed using samtools mpileup. For phylogenetic analysis, site statistics were generated using samtools mpileup and sites filtered using the following criteria: mapping quality (MQ) above 30; site quality score (QUAL) above 30; at least four reads covering each site (supporting either the reference (REF) or a variant (ALT) base call), with at least two reads mapping to each strand; at least 75% of reads supporting site (DP4); allelic frequency (AF1) of one. Sites which failed these criteria in any isolate were removed from analysis.

### Phylogenetic reconstruction

Phylogenetic reconstruction was performed using RAxML (v8.2.3) [31] with a GTR model of nucleotide substitution and a GAMMA model of rate heterogeneity; branch support values determined using 1000 bootstrap replicates. Phylogenetic placement of MinION sequence data was performed by generating site statistics as above, followed by tree estimation using RAxML. Single nucleotide polymorphism (SNP) distance, defined as the number of SNP differences between the MinION isolate at particular depth and the corresponding isolate on the Illumina tree was obtained using python package ete3 [32].

### Sequencing accuracy of MinION

MinION sequencing accuracy at varying depth of coverage was assessed by comparing MiSeq and MinION variant calling across 68 nucleotide positions that contribute to 37 well-characterised non-plasmid resistance associated markers (RAMs) for penicillin, ciprofloxacin, azithromycin and tetracycline resistance (Table 2) [8, 16, 33]. Isolates with MiSeq sequence depth of <30x, our ‘gold standard’, were excluded from analysis, as were

**Table 2.**
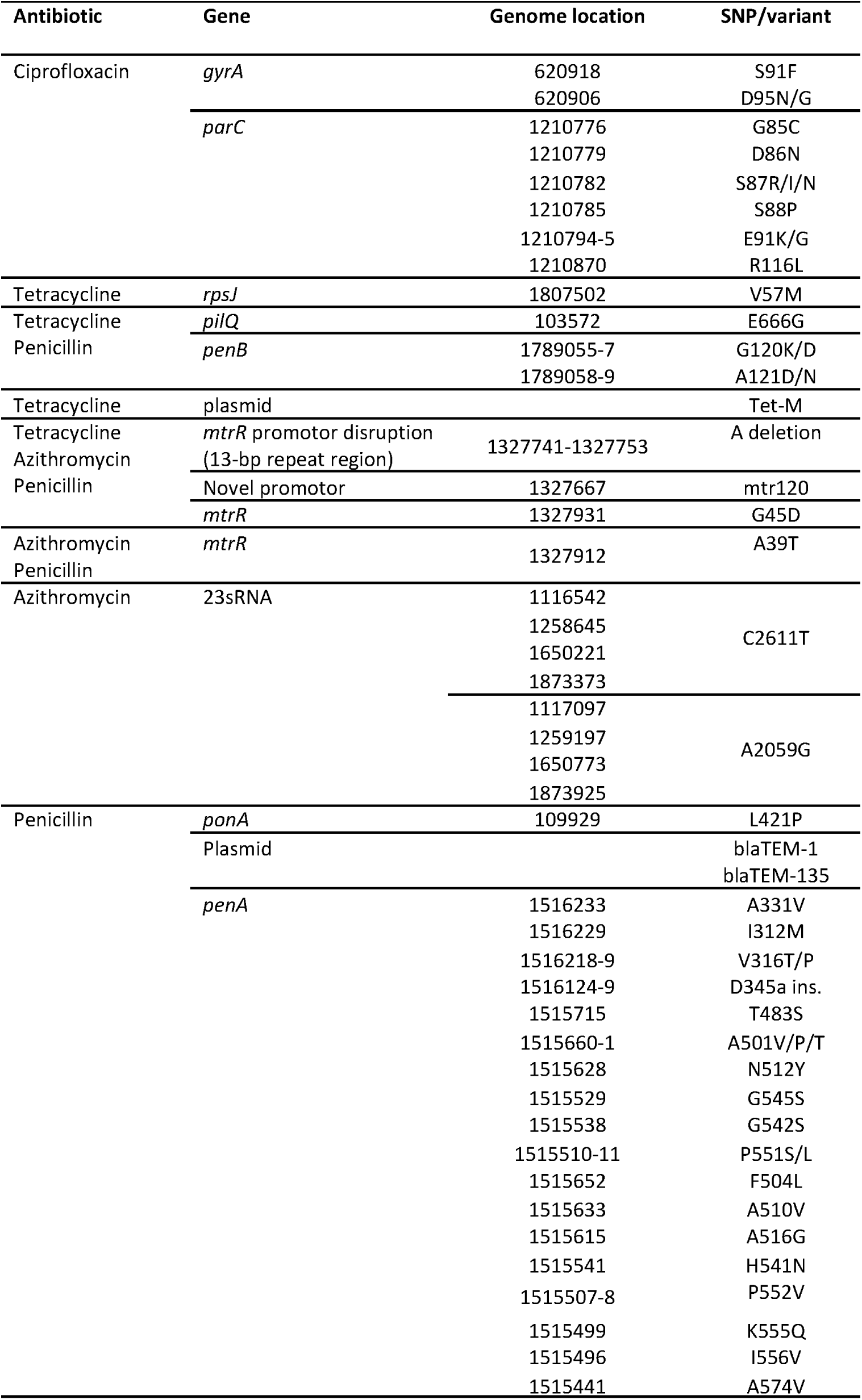
Resistance Associated Markers. Characterised resistance associated markers (RAMs) associated with ciprofloxacin, azithromycin, tetracycline and penicillin resistance in *Neisseria gonorrhoeae* (37 non-plasmid, and three plasmid). Gene name, genome coordinates given for NC_002946 reference genome and SNP or variation linked to resistance, are detailed.

MinION sequences not reaching a final depth of ≥40x, to ensure unbiased retrospective comparison across different depths. Also excluded were two penicillin plasmids and tetracycline tet-M plasmid, as their presence was expected to be determined near immediately. A ‘required sequencing depth’, defined as a suitable depth above which no significant improvement in accuracy of sequencing was achieved, was calculated as a surrogate for the shortest sequencing time providing accurate results.

### Accuracy of MinION molecular distances between isolates

Using the variable nucleotide positions used to construct the maximum-likelihood phylogenetic tree, SNP distances between pairs of whole genome sequences were classified by estimated yearly evolutionary distance (YED), based on the literature described above. Thus, a distance of ≤5 SNPs reflected a possible transmission pair, or isolates separated by one year of time. Subsequent YEDs were then defined by multiples of five SNPs, therefore: <1 year, 1-<2 years, 2-<3 years, 3-<4 years, 4-<10 years, 10-<50 years and ≥50 years, corresponded to 0-4, 5-9, 10-14, 15-19, 20-49, 50-249 and ≥ 250 SNP distances respectively (Figure 1) [17].

**Figure 1.**
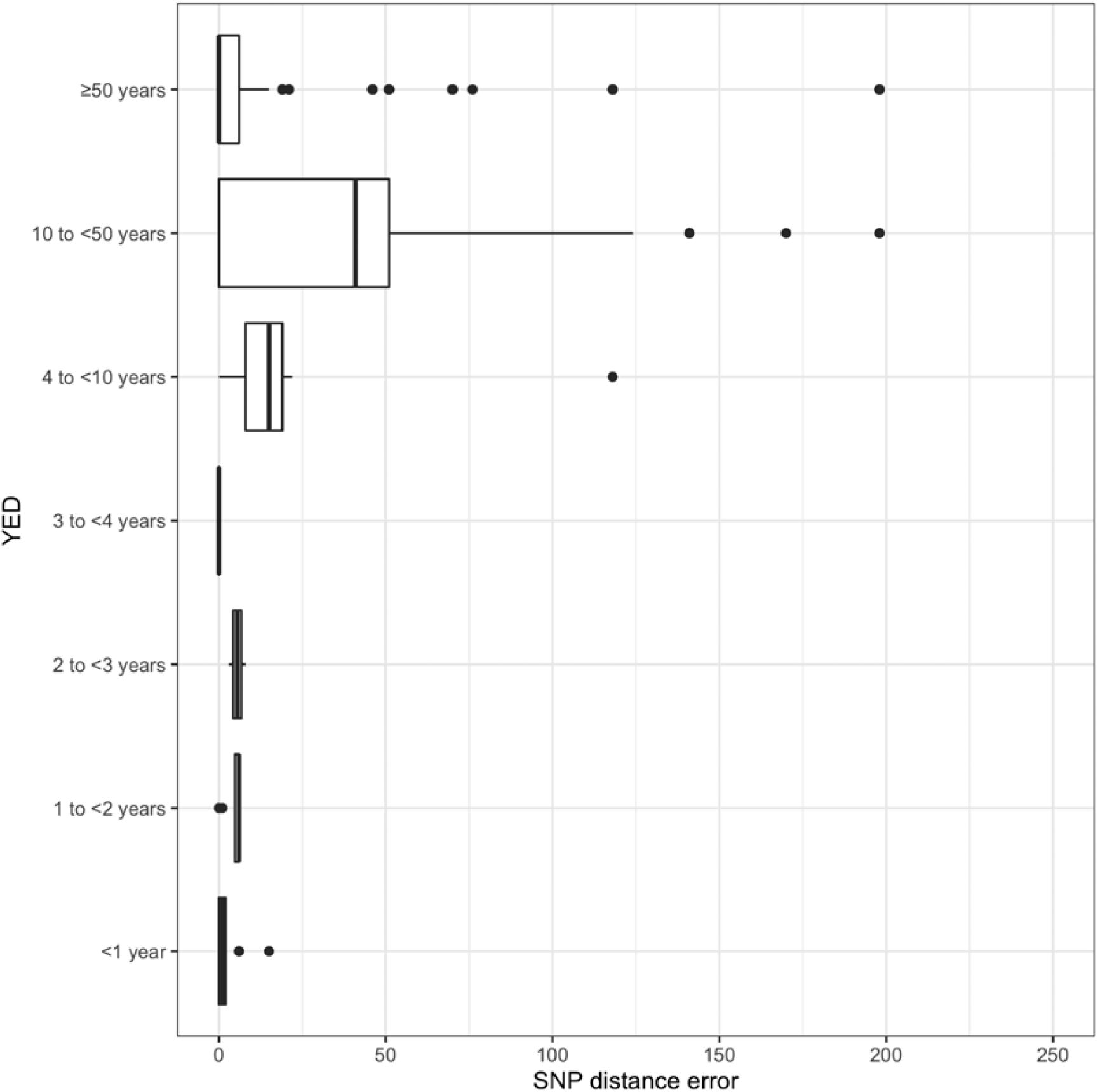
Phylogenetic accuracy of MinION 10x sequencing depth, measured in YEDs. Pairwise SNP distances between MiSeq-MiSeq whole genome sequences grouped by yearly evolutionary distance (YED) (assumption: one YED=5 SNPs). Two corresponding MiSeq-MinION SNP distances (created by substituting each of the isolates in the MiSeq-MiSeq pairs, for its MinION sequence) were compared to the MiSeq-MiSeq SNP distances (of the same isolates). The difference seen by the introduction of a MinION sequence, known as the SNP distance error, is represented by the box and whisker plot. YED count for each category: >50 years - 1848, 10 to <50 years - 66, 4 to <10 years - 18, 3 to <4 years - 2, 2 to <3 years - 2, 1 to <2 years - 8, <1 year - 36.

MinION to MiSeq pairwise SNP distances were calculated by substituting in a MinION sequence for one of each pair, thus creating two MinION-MiSeq distances for each MiSeq-MiSeq distance and isolate pair. The discrepancy in SNP number between the two MinION-MiSeq distances for each MiSeq-MiSeq distance, is defined as the SNP distance error. The YED categories were plotted against MinION-MiSeq SNP distance error, and ANOVA used to determine how the latter varied as YED increased.

### Post-hoc quality control adjustment

A retrospective analysis to determine impact of relaxing variant calling MinION quality control (QC) parameters, for the 68 nucleotide positions, was performed using two alternative QC filters. See supplementary material for detailed methodology.

### Ethics

All examined gonococcal isolates were collected as part of routine diagnostics (standard care) before being anonymised and submitted to GRASP. GRASP is a routine public health surveillance activity, and no specific consent is required from the patients. Patients are informed about GRASP at every participating site, using written notices. The UKHSA has permission to handle data obtained by GRASP under section 251 of the UK National Health Service Act of 2006 (previously section 60 of the Health and Social Care Act of 2001), which was renewed annually by the ethics and confidentiality committee of the National Information Governance Board until 2013. Since then, the power of approval of public health surveillance activity has been granted directly to the UKHSA. Isolates were returned for culture and sequencing with only sample site and gender details used, therefore ethics approval was not needed.

## Results

### N. gonorrhoeae isolate characteristics

Of 58 isolates grown and subsequently sequenced on MiSeq, 47 were successfully re-grown, and sequenced on MinION. Two further isolates were excluded from analysis, one due to no MIC data being available, and another following analysis which suggested the MiSeq and MinION sequences came from different isolates, likely due to laboratory error. Thus MinION, MiSeq and MIC data were initially available for 45 confirmed *N. gonorrhoeae* isolates (Table 1).

### MiSeq and MinION sequencing

Sequence characteristics for individual isolates including median length of *N. gonorrhoeae* mapped reads and final depth of coverage achieved by MiSeq and MinION in all 45 isolates are detailed in Supplementary Table 1. Phylogenetic reconstruction of MiSeq sequences enabled assessment of isolate diversity. Reference alignments were used to identify presence of known RAMs, and phenotype and sample sites were overlaid on the reconstruction (Figure 2). The relationship between sequencing depth and sequencing time is shown in Supplementary Figure 1.

**Figure 2.**
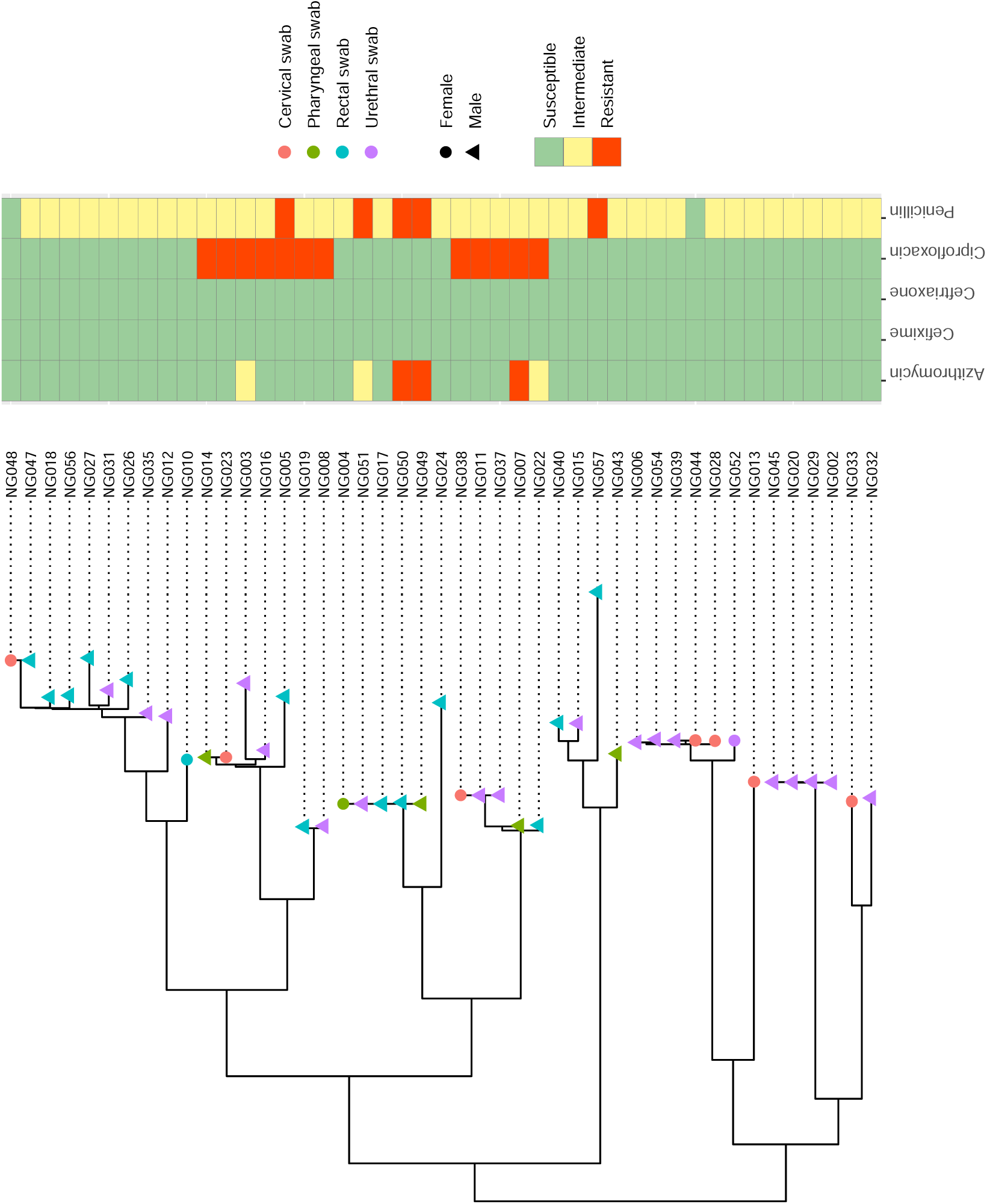
Phylogenetic tree of MiSeq whole genome sequences. Phylogenetic and antimicrobial susceptibility/resistance comparisons of 45 gonococcal isolates. Node shapes represent gender of original sample source, and color sample site. Phenotypic susceptibility profiles are shown as a heatmap as susceptible (green), intermediate (yellow) and resistant (red).

### Genotypic prediction of resistance

The ability to identify known non-plasmid RAMs was assessed for 68 nucleotide positions in 22 isolate MiSeq and MinION sequence pairs, where MinION had reached a final depth of ≥40x. Retrospective analysis of these 22 isolate sequences at 10x, 30x and 40x depth revealed the median number of 68 possible positions per isolate passing standard MinION QC was low at 8 (IQR:5-11), 23 (IQR:18-25) and 25 (IQR:22-30) positions respectively. Combining all nucleotide positions that did pass MinION QC from all 22 isolate sequences, overall MiSeq variant call agreement was 100% (185/185, 95%CI:98.0-100.0), 99.8% (502/503, 95%CI:98.9-99.9) and 99.8% (564/565, 95%CI:99.0-100.0) at 10x, 30x and 40x MinION depth, respectively (Supplementary Table 2).

QC parameters for the 22 MinION isolate sequences at 10x depth were not passed for a nucleotide position within codon 91 of GyrA (genome nucleotide position 620918). However, at 30x and 40x depth, 10 and 12 of the 22 isolates respectively passed QC parameters, and of those 100% matched the sequence determined by MiSeq at the corresponding position, which included both REF and ALT calls. However, four of the 22 isolates that failed QC for a nucleotide in codon 91 at 10x depth and 13 at both 30x and 40x depth, all passed QC at a variant position within codon 95 of GyrA (genome nucleotide position 620906), with 100% agreement to the MiSeq data (Supplementary Table 2C). In this position, all calls passing QC accurately were REF calls only.

In addition, there were insufficient high quality variant calls covering the 13-bp repeat region, within the *mtrR* promotor region, to identify a single base deletion in the 5-A repeat region, present in five of the 22 isolates, based on the MiSeq variant calling results.

### Identification of sequence clusters

A maximum-likelihood phylogenetic tree was constructed, using 7238 variable nucleotide positions present in consensus MiSeq sequences in all 45 isolates that met sequencing QC parameters (Figure 2). Accuracy of placement of individual MinION consensus sequences on to the MiSeq tree was assessed by measuring the MiSeq-MinION pairwise SNP distance of the same isolates. Figure 3 shows the SNP distances over increasing average MinION sequencing depth. SNP distances reduced considerably up to a sequencing depth of 15x (median SNP distances: depths 9x:0.70, 10x:0.20, 15x:0.08). Based on this and the increased sequencing time needed to achieve 15x depth of coverage, we selected 10x as the required sequencing depth. A depth of 10x corresponded to a median of 29 minutes (IQR:25–39 minutes) sequencing time (Supplementary Figure 1). For the 45 isolates, SNP distances between all MiSeq pairs (45×44; n=1980), not including self-comparison, ranged from 0–4053 SNPs. Following categorization of SNP distances between pairs of whole genome sequences into YEDs, most pairs were separated by more than 50 years of molecular distance, and 18 pairs were ‘closely’ related within one YED. For each MiSeq-MiSeq pair, two MinION-MiSeq SNP distances were calculated.

**Figure 3.**
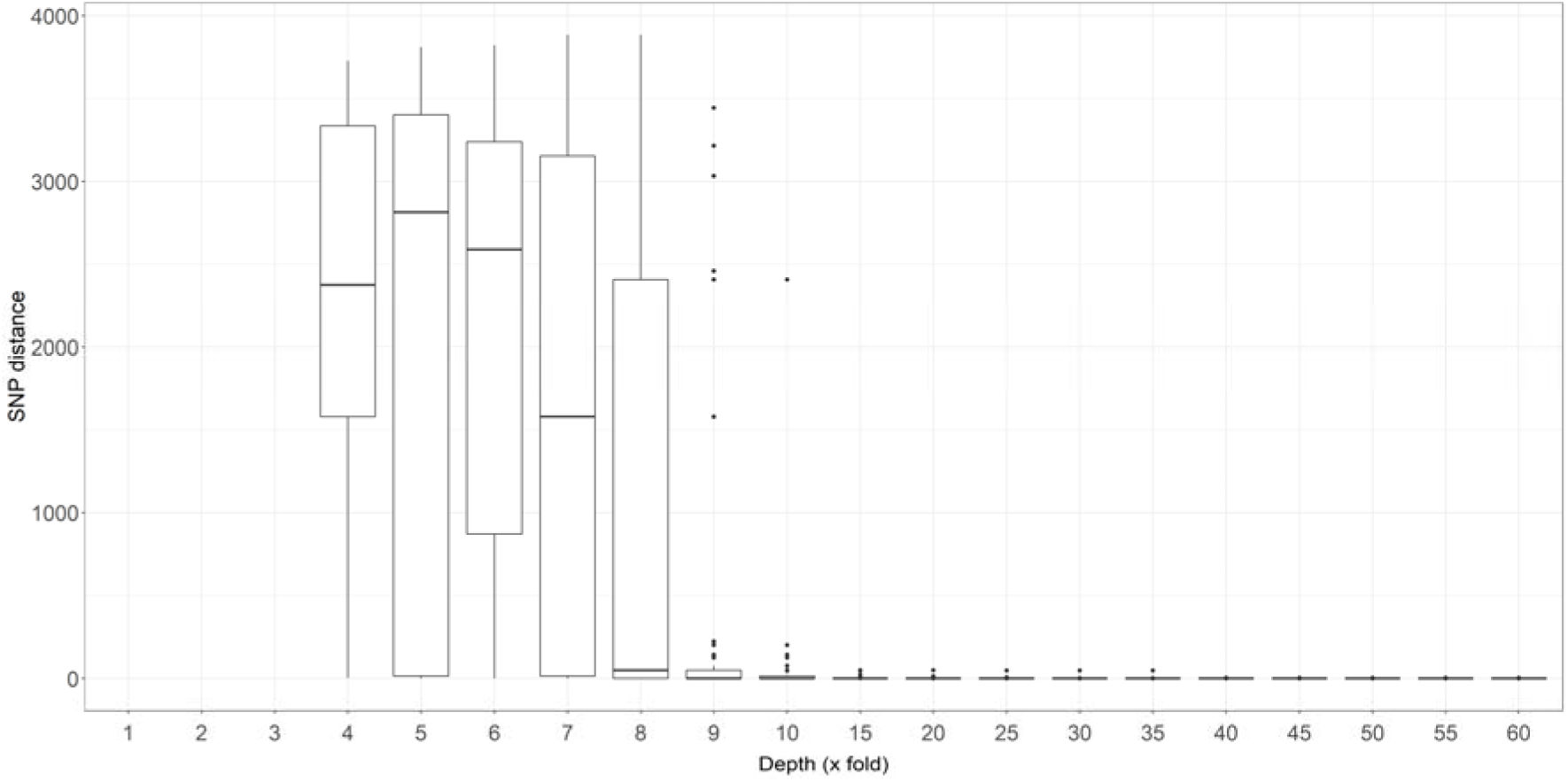
SNP distances between isolate MinION and MiSeq sequences. Box and whisker plot representing Median and IQR of SNP distances between MinION and MiSeq sequences of the same isolate, at increasing MinION sequencing depth of coverage.

Although median SNP distance error in pairwise distance between MinION-MiSeq and MiSeq-MiSeq sequences were similar across all YED categories: 1 (IQR:0-2), 6 (IQR:5-6), 6 (IQR:4-7), 0 (IQR:0-0), 15 (IQR:8-19), 41 (IQR:0-51), and 0 (IQR:0-6), for <1 year, 1-<2 years, 2-<3 years, 3-<4 years, 4-<10 years, 10-<50 years and ≥50 years, respectively (p=0.913, Figure 1), MinION-MiSeq SNP distance errors of >50 were seen only in YED categories 10-<50 and ≥50. Within YED category of <1 year, there were six isolate pairs with a SNP distance error of six, and two of 15.

### Post-hoc quality control adjustment

A post-hoc exploration into low MinION QC rates in RAMs suggested ‘stringent’ QC parameters used, possibly played a part in several variant calls being missed. The largest proportion of all variant calls failing QC did so due to insufficient MQ and DP4 values, particularly due to so called strand-bias. As expected, relaxing QC parameters increased pass rates (Supplementary Table 2). See supplementary material for detailed results.

## Discussion

Our study suggests that nanopore sequencing can accurately perform rapid, flexible and real-time identification of closely related gonococcal strains, offering potential to detect transmission networks. Concerns around drug resistant gonorrhoea particularly to extended spectrum cephalosporins [34], suggest evaluations of established and novel surveillance approaches in/near clinic are warranted. The low cost and portability of the MinION platform, facilitates rapid sequencing in remote areas, with the development of methods for sequencing directly from samples, enabling the use of sequencing information in relative real time with consumable costs likely to become affordable for routine use [35].

As expected, nanopore sequencing accuracy as compared with MiSeq increased progressively with increasing depth. However, as little as 30 minutes of nanopore sequencing, equivalent to 10x average sequencing depth identified closely related gonococcal strains determined by MiSeq sequencing as our gold standard comparison, where final read depths were >30x. We chose a conservative estimate of five SNPs to represent an evolutionary distance of one year between strains, with previous studies suggesting such a distance may also be applicable to determining likely transmission pairs [17, 20].

We were unable to directly compare MinION and MiSeq phylogenetic tree topologies, as too few common SNPs among the MinION sequences passed quality-control. Instead, we calculated SNP distances between every possible pair of MiSeq sequences, categorised them into yearly evolutionary distances, and compared them to MinION-MiSeq SNP distances for the same pairs (i.e., two distances per pair). MinION sequencing at 10x depth accurately measured 28 of the 36 MinION-MiSeq distances, representing the 18 pairs of isolates that fell within a molecular distance of less than one year, with a further two distances falling within a less conservative measure for a year of molecular clock [17]. None of the pairs categorised by MiSeq-MiSeq as greater than one year, were incorrectly assigned to the <1 YED category by the MinION. Although, statistically there was no difference between SNP distance errors across YED categories, more outliers were seen with increasing yearly distance, a reflection of the increasing variant number and low read depth used.

This study was not designed to demonstrate accuracy of predicting AMR phenotype for multiple classes of antibiotics, due to the complexity of having multiple determinants for some antibiotic classes [36]. We did however evaluate the ability to correctly call specific RAMs associated with AMR, which were comparable at 10x, 30x and 40x MinION depth. Previous work has demonstrated a WGS-based MIC prediction model which successfully predicted MIC values with 93% accuracy within +/- 1 doubling dilution and may be adaptable to lower sequencing depths [16]. The AMR RAMs chosen for this analysis were not an exhaustive list and were selected for being well characterised.

Despite the accuracy of rapid nanopore sequencing for specific RAMs being high overall, the large number of positions that did not pass standard QC parameters would not allow for this version of the technology to be used for AMR diagnostic purposes, however this is likely to have changed with improved versions of the platform and flow cell technology. Some RAMs, such as the deletion of a single adenine in the *mtrR* promotor region, were not detected due to poor QC pass rate and the challenges for MinION handling of homopolymers. However, more recent versions of the platform have addressed this problem to some degree [37].

The high QC failure rate prompted post-hoc re-analysis with less stringent QC criteria for the RAMs. These increased the number of RAMs that could be evaluated without losing accuracy, except in the case of the least stringent QC parameters, ‘filter 2’. However, QC rates were still high, exemplified by the nucleotide positions corresponding to the S91 GyrA codon position, a high confidence marker for ciprofloxacin susceptibility, where at 10x depth the number of isolates passing QC at this position increased from zero to seven, with 100% accuracy.

This study used gonococcal isolates and did not perform bioinformatics analysis in real time. Future approaches will likely need to use techniques such as DNA capture to facilitate sequencing directly from samples [22] or more general metagenomic approaches [26] and remove the need for culture, to reduce overall time taken. Automated bioinformatic pipelines will also enable data interrogation in real-time, e.g. the ARTIC network RAMPART (Read Assignment, Mapping, and Phylogenetic Analysis in Real Time) software, a bioinformatics protocol for the analysis of Nanopore sequences [38].

Nanopore technology is undergoing continuous evolution, upgrading the performance of its sequencing technology via improved accuracy in base-calling algorithms, increased throughput and capture, and better raw read accuracy [39]. Additionally, newer rapid library preparation methods may also contribute to reducing the time taken from sample to sequence. The ‘flongle’ flow cell is smaller and cheaper than the conventional MinION flow cells, and adaptable to lower sample number applications such as this. The more recent R10 nanopore further improves the accuracy for detecting homopolymers, and the transition from HMM-based approach to a deep learning approach have improved the read accuracy dramatically [37]. Research is ongoing into sequencing directly from clinical samples, with Street *et al* reporting successful extraction and sequencing of *N. gonorrhoeae* DNA directly from urine samples, achieving a coverage of ≥92.8% across the whole genome in ten patient samples, and ≥93.8% at ≥10x sequencing depth in seven [22]. The combination of these improvements suggest that MinION offers promise as a tool to widen rapid *N. gonorrhoeae* surveillance for monitoring AMR spread.

In conclusion, MinION sequencing was able to accurately determine closely related gonococcal strains, related within a molecular distance of one year, with as little as 10x average depth of coverage, demonstrating the potential for its application as a real-time surveillance tool.

## Data Availability

Anyone can share this material, providing it remains unaltered in any way and the original authors are credited and cited.

https://www.ebi.ac.uk/ena/browser/view/PRJEB50649?show=reads

## Acknowledgements

We thank the staff and patients of The Courtyard Clinic, St George’s Healthcare NHS Trust. We also thank the clinical and laboratory staff at the UKHSA who contributed to the Gonococcal Resistance to Antimicrobials Surveillance Programme (GRASP), for returning the isolates used in this study.

