## Supplementary Material for "Speed and accuracy of whole-genome nanopore sequencing for differential *Neisseria gonorrhoeae* strain detection in samples collected prospectively from a sexual health clinic"

**Methods**

**MinION library preparation**: Where possible 1 µg, or the maximum amount available up to 1 µg, of genomic DNA of each of the 47 samples was dissolved in 50 µl of nuclease-free water and fragmented using a Covaris g-TUBE (Covaris, USA) according to manufacturer's instructions (centrifuged at 6000 rpm for 60 seconds, in both directions). To confirm fragmentation, 1 µl of each sample DNA was run on the 2100 Bioanalyzer instrument (Agilent Technologies, USA). End repair and dA tailing were performed on the sheared DNA using NEBNext® Ultra™ II End repair/dA Tailing Module (NEB, UK) and end-prepped DNA purified using solid phase reversible immobilisation (SPRI) purification (Beckman Coulter, UK), eluting in 22.5 µl of nuclease-free water according to manufacturer's instructions. Twelve native barcodes NB01-NB12 from kit EXP-NBD103 (ONT, UK) were ligated to 12 samples of end-prepped DNA, using 22.5 µl end-prepped DNA, 2.5 µl of native barcode adaptor and 25 µl of Blunt TA master mix (NEB, UK). A second SPRI purification was carried out according to manufacturer's instructions and DNA eluted in 15 µl with water. DNA was quantified using Qubit v1.0 with Qubit dsDNA BR assay kit (Invitrogen, USA). Equal amounts of each sample were pooled into sets of three, with a total pooling mass of approximately 700 ng, equalling approximately 233 ng per sample. 20 µl of native barcoding adapter mix (BAM) was ligated to each pool using 50 µl Blunt TA master mix, and made up to 100 µl with water. A final SPRI purification step was done using 40 µl of SPRI beads, followed by 2 x 140 µl wash with adapter bead binding (ABB) and elution in 25 µl of elution buffer (ELB). The library was then ready for sequencing on the MinION.

MinION sequencing was performed using 16 MinION flow cells (Flow Cell - FLO-MIN105 using R9.2 Pore) with barcoded-DNA from three isolates per flow cell. Each MinION flow cell was removed from its packaging and inserted onto the MinION 1.1b (MIN101B) and MinION quality control software run to assess pore activity. Barcoded one-directional DNA libraries were created using library kits: SQK-NSK007 and EXP-NBD103, and 12 µl of each barcoded sample pool was loaded into the MinION according to manufacturer's instructions. Flow cells were run for varying lengths of time, between 2–24 hours. No flow cells were reused. 1D base calling was performed using the Metrichor rev 1.107 (cloud) platform.

**Post-hoc quality control adjustment:** The first, 'filter 1', maintained all QC parameters of standard QC (as detailed in the methods), with alteration to only the DP4 filtering criteria to include at least one read mapping to each strand (forward and reverse) of either the REF or ALT site, and with these minimum criteria met, at least 75% of total number of reads supporting that site. The second, 'filter 2', also maintained all QC parameters of standard QC with a further relaxation of DP4 criteria to a minimum of only one read covering one strand of either the REF or ALT site, and with these minimum criteria met, at least 75% of reads supporting that site. Additionally, the impact on accuracy when compared to the MiSeq position within the same isolate, was done.

### Results

**Post-hoc quality control adjustment:** For standard QC parameters, DP4 requires a minimum of four reads, with at least two mapping to each strand of one site (supporting the REF or ALT), and at least 75% of reads supporting that site. In many cases, strand-bias was caused by sufficient reads for only one strand (on one site), and less than two for the opposite strand, resulting in QC failure.

The relaxation of QC parameters using filter 1 and filter 2, increased QC pass rates with increasing depth of coverage (Supplementary Table 2). Overall, increased QC pass rate was not associated with loss of accuracy. The impact of filter 1 QC parameters on genome position 620918 within codon 91 in GyrA, was an increase in number of positions passing QC, as well as maintaining accuracy at 100% when compared to the MiSeq, across all three depths. For genome position 620906 within codon 95 of GyrA, filter 1 QC parameters saw the number that passed QC increase from 4/22 to 9/22 at 10x depth, however no further increase was seen at 30x and 40x depth. All sites that passed filter 1 QC at this position agreed with the MiSeq and maintained 100% accuracy. Filter 2 QC parameters, saw an increase in isolate numbers at both positions in GyrA that passed QC at depths of 10x, 30x and 40x, but with decreased accuracy (Supplementary Table 2, C & D).

**Supplementary Table 1. Summary of MinION and MiSeq sequencing outputs**

| Isolate | MiSeq | | MinION | | |
|---|---|---|---|---|---|
| | Read count | Depth | Read count | Depth | Median read length |
| NG002 | 286726 | 59 | 37791 | 95 | 5228 |
| NG003 | 374461 | 80 | 32921 | 89 | 5906 |
| NG004 | 359432 | 79 | 32297 | 80 | 4724 |
| NG005 | 398056 | 84 | 16470 | 42 | 5268 |
| NG006 | 329834 | 75 | 66837 | 80 | 2013 |
| NG007 | 382478 | 79 | 37944 | 82 | 4477 |
| NG008 | 144410 | 26 | 58890 | 76 | 1957 |
| NG010 | 181701 | 41 | 28766 | 50 | 3489 |
| NG011 | 254192 | 55 | 21676 | 40 | 4016 |
| NG012 | 235162 | 54 | 29130 | 39 | 2491 |
| NG013 | 125864 | 22 | 37164 | 75 | 4311 |
| NG014 | 249449 | 54 | 36783 | 57 | 3096 |
| NG015 | 223409 | 46 | 19411 | 47 | 5598 |
| NG016 | 209404 | 45 | 20056 | 41 | 3802 |
| NG017 | 142738 | 30 | 21572 | 39 | 3882 |
| NG018 | 186799 | 42 | 40262 | 90 | 4891 |
| NG019 | 287033 | 63 | 32981 | 68 | 4416 |
| NG020 | 279821 | 60 | 84971 | 105 | 1813 |
| NG022 | 251790 | 53 | 39907 | 65 | 3409 |
| NG023 | 276267 | 62 | 57729 | 80 | 2488 |
| NG024 | 354985 | 75 | 26719 | 59 | 4767 |
| NG026 | 240317 | 54 | 19265 | 43 | 4865 |
| NG027 | 283617 | 61 | 26274 | 54 | 4487 |
| NG028 | 204630 | 42 | 36560 | 51 | 2559 |
| NG029 | 121248 | 21 | 13602 | 30 | 5154 |
| NG031 | 273662 | 63 | 13538 | 29 | 4489 |
| NG032 | 326198 | 68 | 5117 | 11 | 4299 |
| NG033 | 164870 | 37 | 21286 | 24 | 1883 |
| NG035 | 185770 | 38 | 18782 | 26 | 2131 |
| NG037 | 274239 | 61 | 17889 | 26 | 1658 |
| NG038 | 197600 | 41 | 27064 | 34 | 1755 |
| NG039 | 241124 | 54 | 15144 | 19 | 1774 |
| NG040 | 237790 | 51 | 8086 | 19 | 5672 |
| NG043 | 169118 | 28 | 80153 | 67 | 1355 |
| NG044 | 220790 | 45 | 27641 | 32 | 1636 |
| NG045 | 190776 | 43 | 26785 | 36 | 2043 |
| NG047 | 173802 | 39 | 32648 | 34 | 1412 |
| NG048 | 253662 | 56 | 25489 | 31 | 1576 |
| NG049 | 88812 | 17 | 39523 | 40 | 1499 |
| NG050 | 34598 | 6 | 45642 | 46 | 1188 |
| NG051 | 382287 | 83 | 24154 | 32 | 2067 |
| NG052 | 510395 | 111 | 49371 | 42 | 1500 |
| NG054 | 275550 | 58 | 47080 | 40 | 1427 |
| NG056 | 314907 | 71 | 13558 | 32 | 5172 |
| NG057 | 169721 | 31 | 15576 | 32 | 4892 |

Read count and depth of coverage of MinION and MiSeq sequencing, with median read length for MinION only. Sequencing depth was calculated based on reads mapped against the FA1090 reference genome (accession: NC_002946).

**Supplementary Table 2. Number and accuracy of MinION variant calls passing standard and alternative QC parameters**

**A.**

| **Passed QC** | Standard QC | Filter1 | Filter2 |
|---|---|---|---|
| 10x | 13.5% | 28.7% | 52.8% |
| | 185/1371 | 393/1372 | 724/1371 |
| 30x | 36.5% | 47.9% | 70.2% |
| | 503/1378 | 660/1378 | 967/1378 |
| 40x | 40.9% | 51.3% | 72.7% |
| | 565/1382 | 709/1382 | 1005/1382 |

**B.**

| **Passed QC + accuracy** | Standard QC | Filter1 | Filter2 |
|---|---|---|---|
| 10x | 100% | 100% | 99.2% |
| | 185/185 | 393/393 | 718/724 |
| 30x | 99.8% | 99.8% | 98.8% |
| | 502/503 | 659/660 | 955/967 |
| 40x | 99.8% | 99.7% | 98.8% |
| | 564/565 | 707/709 | 993/1005 |

**C.**

Position: 620906 (GyrA D95)

| **Passed QC + accuracy** | Standard QC | Filter1 | Filter2 |
|---|---|---|---|
| 10x | 100% | 100% | 86.7% |
| | 4/4 | 9/9 | 13/15 |
| 30x | 100% | 100% | 81.3% |
| | 13/13 | 13/13 | 13/16 |
| 40x | 100% | 100% | 81.3% |
| | 13/13 | 13/13 | 13/16 |

Position: 620918 (GyrA S91)

| **Passed QC + accuracy** | Standard QC | Filter1 | Filter2 |
|---|---|---|---|
| 10x | 0 | 100% | 100% |
| | 0/0 | 7/7 | 13/13 |
| 30x | 100% | 100% | 100% |
| | 10/10 | 14/14 | 16/16 |
| 40x | 100% | 100% | 100% |
| | 12/12 | 14/14 | 17/17 |

Total number of the 68 nucleotide positions representing 37 non-plasmid RAMs across 22 MinION isolates which: **A**. passed standard, filter 1, and filter 2 QC parameters **B**. passed standard, filter 1, and filter 2 QC parameters, and were accurate when compared to the MiSeq sequences within the same isolate, at the same position **C**. Total number of positions: 620906, and 620918 corresponding to GyrA D95 and S91 respectively, that passed standard, filter 1, and filter 2 QC parameters, and were accurate when compared to the MiSeq sequences within the same isolate, at the same position.

**Supplementary Figure 1. MinION genome depth of coverage vs sequencing time**

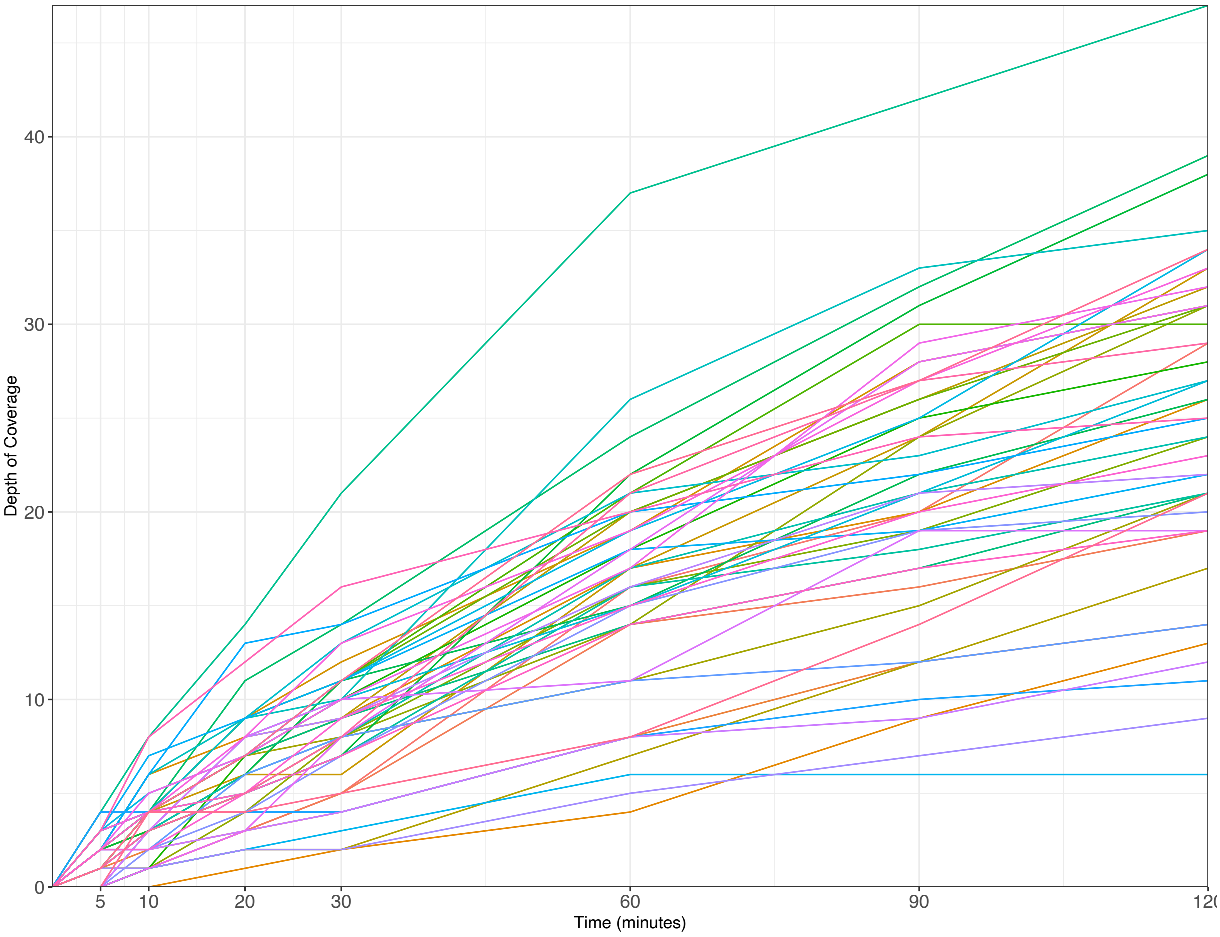


MinION depth of coverage at increasing timepoints.
